# Beyond Algorithms: The Impact of Simplified CNN Models and Multifactorial Influences on Radiological Image Analysis

**DOI:** 10.1101/2024.09.15.24313585

**Authors:** Saber Mohammadi, Abhinita S. Mohanty, Shady Saikali, Doori Rose, WintPyae LynnHtaik, Raecine Greaves, Tassadit Lounes, Eshaan Haque, Aashi Hirani, Javad Zahiri, Iman Dehzangi, Vipul Patel, Pegah Khosravi

## Abstract

This paper demonstrates that simplified Convolutional Neural Network (CNN) models can outperform traditional complex architectures, such as VGG-16, in the analysis of radiological images, particularly in datasets with fewer samples. We introduce two adopted CNN architectures, LightCnnRad and DepthNet, designed to optimize computational efficiency while maintaining high performance. These models were applied to nine radiological image datasets, both public and in-house, including MRI, CT, X-ray, and Ultrasound, to evaluate their robustness and generalizability. Our results show that these models achieve competitive accuracy with lower computational costs and resource requirements. This finding underscores the potential of streamlined models in clinical settings, offering an effective and efficient alternative for radiological image analysis. The implications for medical diagnostics are significant, suggesting that simpler, more efficient algorithms can deliver better performance, challenging the prevailing reliance on transfer learning and complex models. The complete codebase and detailed architecture of the LightCnnRad and DepthNet, along with step-by-step instructions, are accessible in our GitHub repository at https://github.com/PKhosravi-CityTech/LightCNNRad-DepthNet.

## Introduction

Recent advancements in deep learning have significantly impacted medical image analysis, with Convolutional Neural Networks (CNNs) playing a pivotal role in improving the accuracy and efficiency of radiological interpretations. CNNs are widely recognized for their ability to automatically extract relevant features from various medical images, including those acquired through Ultrasound (US) [1], Magnetic Resonance Imaging (MRI) [2], Computed Tomography (CT) [3], and Positron Emission Tomography (PET) [4].

CNNs, a type of deep neural network (DNN) architecture, are particularly adept at processing structured data, such as medical images. They effectively capture both low- and high-level features within radiological images, offering improved efficiency over conventional methods. Typically, CNNs consist of three fundamental building blocks: convolution, pooling, and fully connected layers. The convolution and pooling layers are crucial for extracting hierarchical features, allowing the network to progressively refine its understanding of the input data. Through a combination of convolution and pooling operations, CNNs can adaptively learn representations of medical images and enhance the model’s receptive field within the deeper layers. The feature maps generated by the convolution and pooling layers are then fed into fully connected layers, which are responsible for performing classification tasks [5].

CNNs have emerged as versatile tools in radiology research, serving multiple purposes such as anomaly detection [6], classification [7,8], segmentation [9], and image reconstruction [10,11]. For instance, Khosravi et al. [8] demonstrated the efficacy of their deep learning CNN (DLCNN) model in classifying benign versus malignant tumors and high-risk versus low-risk cases in prostate cancer using MRI scans, achieving remarkable performance (AUC=0.86) [8]. Similarly, Salama et al. [7] utilized various pre-trained DLCNN models for breast cancer classification in mammogram images, achieving an outstanding accuracy and AUC of nearly 90% [7]. The remarkable performance of CNNs, sometimes surpassing that of human experts, underscores their potential in medical image analysis and diagnosis [12]. However, developing and training deep CNN models capable of achieving high accuracy and generalizability in classifying medical images presents a significant challenge. The complexity of very deep CNNs, while advantageous for image classification, increases the risk of overfitting [13]. Additionally, the training process for deep CNNs can be time-consuming, especially when computational resources are limited [13].

This paper introduces two novel CNN architectures—LightCnnRad and DepthNet— specifically designed for radiological applications. These architectures are distinguished by their reduced parameter counts, which facilitate more efficient training compared to more complex models. Additionally, their versatility allows them to be applied across a range of medical imaging modalities, from MRI to CT scans, demonstrating their adaptability to diverse diagnostic needs. The first aim of this study is to address the challenge of high computational costs by proposing efficient models suitable for various clinical environments. The second aim is to emphasize the importance of a holistic approach in radiological image analysis. Despite the high performance of many existing algorithms, optimal results are often compromised by inadequate preprocessing and external factors. This study advocates for a comprehensive approach that goes beyond algorithmic sophistication to include these critical elements. By addressing these two key goals, our research provides essential insights into the multifaceted nature of medical image analysis, guiding future research in this vital domain.

## Materials and Methods

### Datasets

To evaluate the performance of LightCnnRad and DepthNet, we used a combination of eight public datasets and one in-house dataset. The public datasets, sourced from repositories such as Kaggle and The Cancer Imaging Archive (TCIA), encompass various imaging modalities, including MRI, CT, X-ray, and Ultrasound (US). Each dataset is focused on different diseases and imaging techniques, providing a diverse and comprehensive foundation for our analysis. The datasets include MRI scans for Alzheimer’s Disease, CT scans for COVID-19 and lung cancer, ultrasound images for breast cancer, and X-ray images for pneumonia, tuberculosis, and a combined COVID- 19/pneumonia dataset. These datasets vary in size and class distribution, with Alzheimer’s Disease (4,540 images) divided between non-demented and very mild demented classes, COVID-19 (8,001 images) split evenly between positive and negative cases, and lung cancer (560 images) distinguishing between adenocarcinoma and squamous cell carcinoma. The breast cancer dataset (460 images) is categorized into benign and malignant classes, while the pneumonia dataset (3,179 images) and tuberculosis dataset (1,394 images) separate cases with and without the disease. The combined COVID-19/pneumonia dataset (3,173 images) differentiates between these two conditions. Each dataset was sourced from Kaggle, ensuring a diverse and comprehensive foundation for our analysis (Figure 1 and Table 1).

**Figure 1.**
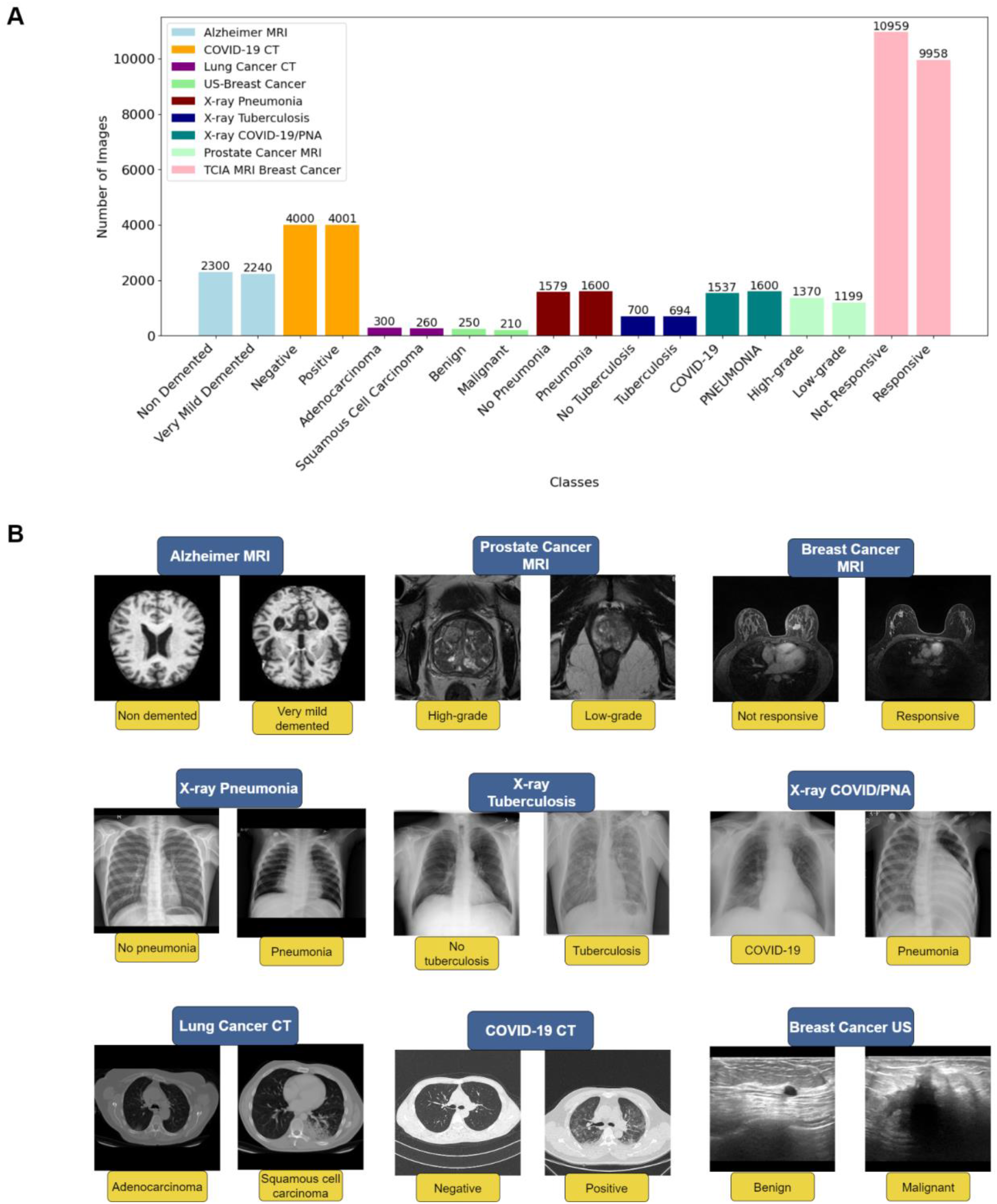
A) The distribution of images across all included datasets. B) Representative sample images from each class within the datasets.

**Table 1.** Number of images and corresponding classes in each dataset.

| Dataset | Imaging Technique | Classes | Total No. of Images | No. of images according to classes | Reference (Link) |
| --- | --- | --- | --- | --- | --- |
| Alzheimer's Disease | MRI | Non Demented Vs. Very mild demented | 4,540 | Nondemented: (n=2,300) | Alzheimer MRI Preprocessed Dataset [16] |
|  |  |  |  | Very mild demented: |  |
|  |  |  |  | (n=2,240) |  |
| COVID-19 | CT | negative vs. positive | 8,001 | Negative: (n=4,000) | CT Scans for COVID-19 Classification [17] |
|  |  |  |  | Positive: (n=4,001) |  |
| Lung Cancer | CT | adenocarcinoma vs. squamous cell carcinoma | 560 | Adenocarcinoma: (n=300) | Chest CT-Scan images Dataset [18] |
|  |  |  |  | Squamous Cell Carcinoma: (n=260) |  |
| Breast Cancer | Ultrasound | benign vs. malignant | 460 | Benign: (n=250) | Breast Ultrasound Images Dataset [19] |
|  |  |  |  | Malignant: (n=210) |  |
| Pneumonia | X-ray | No pneumonia vs. Pneumonia | 3,179 | No pneumonia: (n=1,579) | Chest X-Ray Images (Pneumonia) [20] |
|  |  |  |  | Pneumonia: (n=1,600) |  |
| Tuberculosis | X-ray | No tuberculosis vs. Tuberculosis | 1,394 | No tuberculosis: (n=700) | Tuberculosis (TB) Chest X-ray Database [21] |
|  |  |  |  | Tuberculosis: (n=694) |  |
| COVID-19/PNA | X-ray | COVID-19 vs. Pneumonia | 3,173 | COVID-19: (n=1,537) | COVID19/PNEUMONIA/Normal Chest X-Ray Image Dataset [22] |
|  |  |  |  | Pneumonia: (n=16,00) |  |
| Breast Cancer | MRI | Not responsive vs. Responsive | 20,917 | Not responsive: (n=10,959) | Duke Breast Cancer MRI Dataset [23] |
|  |  |  |  | Responsive: (9,958) |  |
| Prostate Cancer | MRI | High-grade vs. Low-grade (ISUP = 4 and 5 vs. ISUP =1, respectively) | 2,569 | High-grade: (n=1,370) | In-house Dataset |
|  |  |  |  | Low-grade: (n=1,199) |  |

To evaluate the model’s effectiveness in addressing clinically relevant questions, particularly its capability to classify patients based on their therapeutic response, we utilized breast cancer axial MRI images from the Duke-Breast Cancer-MRI dataset, available through The Cancer Imaging Archive (TCIA). This dataset includes axial breast MRI images acquired using 1.5T or 3T scanners from 922 biopsy-confirmed invasive breast cancer patients in the prone position. For this study, we selected pre- and post-contrast images from 278 patients, specifically those with documented complete or near-complete response to neoadjuvant chemotherapy (NAC) and those with no response, to develop a binary classification model (Figure 1 and Table 1). Patients with a complete or near-complete response to NAC were classified as 1 or 2, while those with no response were classified as 0. The analysis was conducted using T1-weighted (T1W) MRI sequences in DICOM format to ensure consistency in image quality and diagnostic relevance. To address the issue of class imbalance, we excluded images from 90 patients in the majority class (non-responsive), resulting in a balanced dataset comprising 188 patients (Train: 131, Test: 29, Validation: 28).

To further assess the performance of our models, we included an in-house dataset of prostate cancer MRI images. The ISUP (International Society of Urological Pathology) grading system for prostate cancer, often referred to as the ISUP 2014/2015 grading system, classifies the aggressiveness of prostate cancer based on histopathological findings. The grades range from 1 to 5 and guide treatment decisions.

For this study, MRI images corresponding to patients with ISUP grade 1 were selected as the low-grade class, while those with grades 4 and 5 were selected as the high-grade class. Our in-house dataset used in this study was specifically curated from preoperative MRI scans of patients with prostate cancer (PCa). The data collection involved a detailed curation and annotation process by fellowship-trained urologists. It includes a comprehensive set of 2,569 T2-weighted (T2W) MR images from 394 patients (Train: 275, Test: 60, Validation: 59), acquired from various vendors between 2018 and 2023. The images, which originated from multiple centers, were standardized to uniform dimensions to ensure consistency and were stored in the hospital PACS system before retrieval for analysis. The dataset is balanced, comprising 1,370 high-grade and 1,199 low-grade images, providing a robust basis for model training (Figure 1 and Table 1). This study was conducted with approval from the AdventHealth Institutional Review Board (IRB) in Orlando, FL, under protocol number 3009855250. The MR images analyzed in this study are not publicly available due to the need for data usage agreements and compliance with privacy and security protocols, given the sensitivity of the medical data. The details of the included datasets are summarized in Table 1.

All datasets were partitioned into training, validation, and test sets following a 70:15:15 ratio. Figure 1A illustrates the distribution of images within each dataset, detailing the proportions assigned to each subset. Additionally, Figure 1B provides representative examples from each class within the datasets.

### Preprocessing

In this study, we applied the same comprehensive preprocessing pipeline to all images, regardless of the specific algorithm used, including LightCnnRad, DepthNet, and VGG-16. VGG-16 [24] was selected for comparison due to its well-established performance in image classification tasks and its ability to serve as a benchmark against newer models like LightCnnRad and DepthNet.

Our preprocessing began with the removal of duplicate images to ensure the uniqueness of each image within the dataset, thereby preventing the models from overfitting due to duplicate patterns. This is particularly crucial for public datasets, where prior preprocessing steps might be unknown. Following this, we converted all image files to the PNG format from their original JPEG/JPG and DICOM formats. PNG was chosen for its lossless compression, preserving image quality and ensuring no critical information was lost during the conversion process. Another crucial step was channel verification, where we confirmed that all images contained three RGB channels, ensuring a standardized input format for all models. We also resized the images to a consistent square dimension, a critical step for batch processing and maintaining uniformity across the dataset. This uniform resizing is particularly important for ensuring that all models can effectively process the images during training. The overall workflow of these preprocessing steps is depicted in Figure 2, illustrating the systematic approach we took to prepare the datasets for optimal model performance across all algorithms employed in the study.

**Figure 2.**
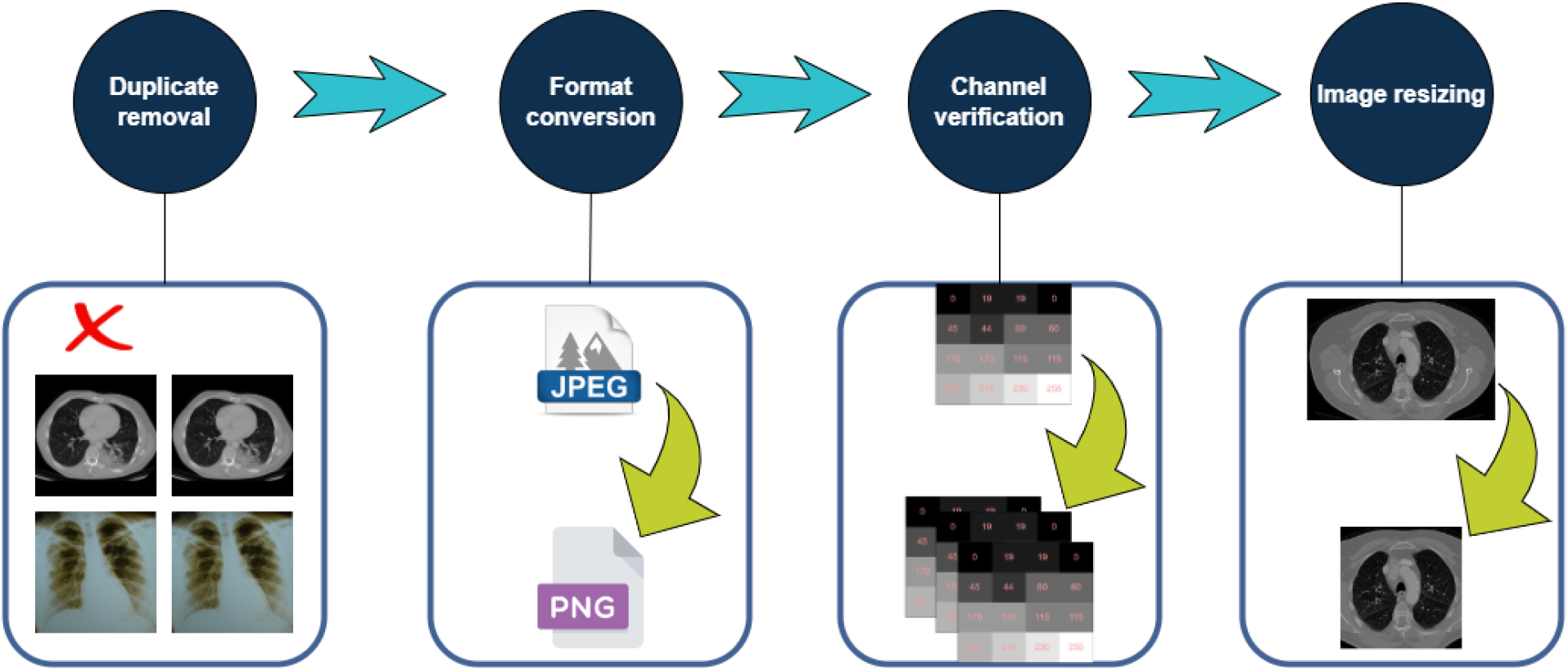
Preprocessing workflow. All datasets underwent comprehensive preprocessing prior to training with LightCnnRad, DepthNet, and VGG-16 models.

While a general preprocessing workflow was maintained for all models, we accommodated specific architectural requirements where necessary. For instance, LightCnnRad and DepthNet required input images of different sizes due to their design. In the case of LightCnnRad, the input images were resized to 150x150 pixels, reflecting the model’s focus on operating efficiently with smaller image dimensions while maintaining classification accuracy. In contrast, DepthNet, which employs a more complex architecture with depthwise separable convolutions, processes images resized to 512x512 pixels. The larger input size is consistent with DepthNet’s need for higher resolution to fully exploit its depthwise separable convolution layers, which are designed to capture more detailed spatial information. These preprocessing differences are tailored to maximize the strengths of each model’s architecture, ensuring that LightCnnRad maintains its efficiency with smaller images while DepthNet leverages higher-resolution inputs for more intricate feature extraction. For all models, the images underwent standard data augmentation through random horizontal flipping, followed by conversion to tensors and normalization across the RGB channels, with pixel values scaled to a range of [-1, 1].

### Training Procedure

LightCnnRad, DepthNet, and VGG-16 were trained using all available datasets on a machine equipped with a 13th Gen Intel Core i7-13650HX processor running at 2.60 GHz, 32 GB of RAM, and an NVIDIA GeForce RTX 4070 GPU. This high-performance setup ensured efficient training and rapid model iterations.

The training process involved running our models for a total of 100 epochs, with early stopping employed to prevent overfitting. We consistently used a learning rate of 0.001 across all datasets, optimized through a grid search to ensure stable and gradual convergence. For LightCnnRad and DepthNet, multiple optimizers were initially tested across different datasets. RMSprop (alpha=0.9 and 0.8, with weight decay=0.0001 and 0.00001 for LightCnnRad and DepthNet, respectively) yielded the best results for most datasets, except for the prostate cancer MRI dataset, where Adam was more effective due to its adaptive learning rate. For VGG-16 models, Stochastic Gradient Descent (SGD, momentum=0.9) provided superior convergence and performance.

Additionally, a batch size of 32 was used, allowing for efficient use of memory and more stable gradient estimates. The cross-entropy loss function was employed to measure the discrepancy between predicted probabilities and true class labels, providing a robust criterion for model optimization. Despite the complexity of classifying the aggressiveness of prostate cancer based on histopathological information, the consistent application of these optimized hyperparameters across all datasets contributed to the stable performance of our models.

Our models were implemented using PyTorch v.2.3 and Cuda v.12.1, which provided a flexible and efficient framework for developing and training deep learning models. PyTorch’s dynamic computational graph and extensive library support facilitated the implementation of our architectures and the execution of the training process. Overall, the training procedure was designed to optimize the performance of our models, ensuring that they can effectively analyze radiological images across various modalities with high accuracy and efficiency.

### The Architecture of LightCnnRad and DepthNet

LightCnnRad and DepthNet are optimized versions of well-established neural network architectures, adapted to balance computational efficiency with high performance in radiological image classification tasks. LightCnnRad, inspired by traditional CNNs, is designed to maintain simplicity while achieving robust classification accuracy. The model features three convolutional layers, each followed by batch normalization and ReLU activation functions. These layers progressively increase in the number of filters (from 12 to 32) and are interspersed with max pooling to reduce spatial dimensions, ultimately producing a compact representation that is passed through a fully connected layer for final classification. This design minimizes computational complexity while preserving high performance, making it suitable for large-scale image analysis.

DepthNet builds upon the architecture of MobileNet [25], employing depthwise separable convolutions to significantly reduce the number of parameters compared to traditional convolutions. This model consists of four blocks, each comprising a depthwise convolution followed by a pointwise convolution, batch normalization, and ReLU activation. These blocks progressively increase the channel dimensions, ensuring detailed feature extraction while maintaining efficiency. The final output is flattened and passed through fully connected layers, with dropout regularization to prevent overfitting, before generating the classification output. By adopting depth-wise separable convolutions, DepthNet achieves a significant reduction in computational demands without compromising on accuracy.

VGG-16 was also selected as a benchmark for comparison due to its well-documented performance in image classification tasks, allowing us to highlight the efficiency and effectiveness of our optimized models, LightCnnRad and DepthNet. Figure 3 (A, B, C, and D) illustrates the architectures and parameter counts for all three models.

**Figure 3.**
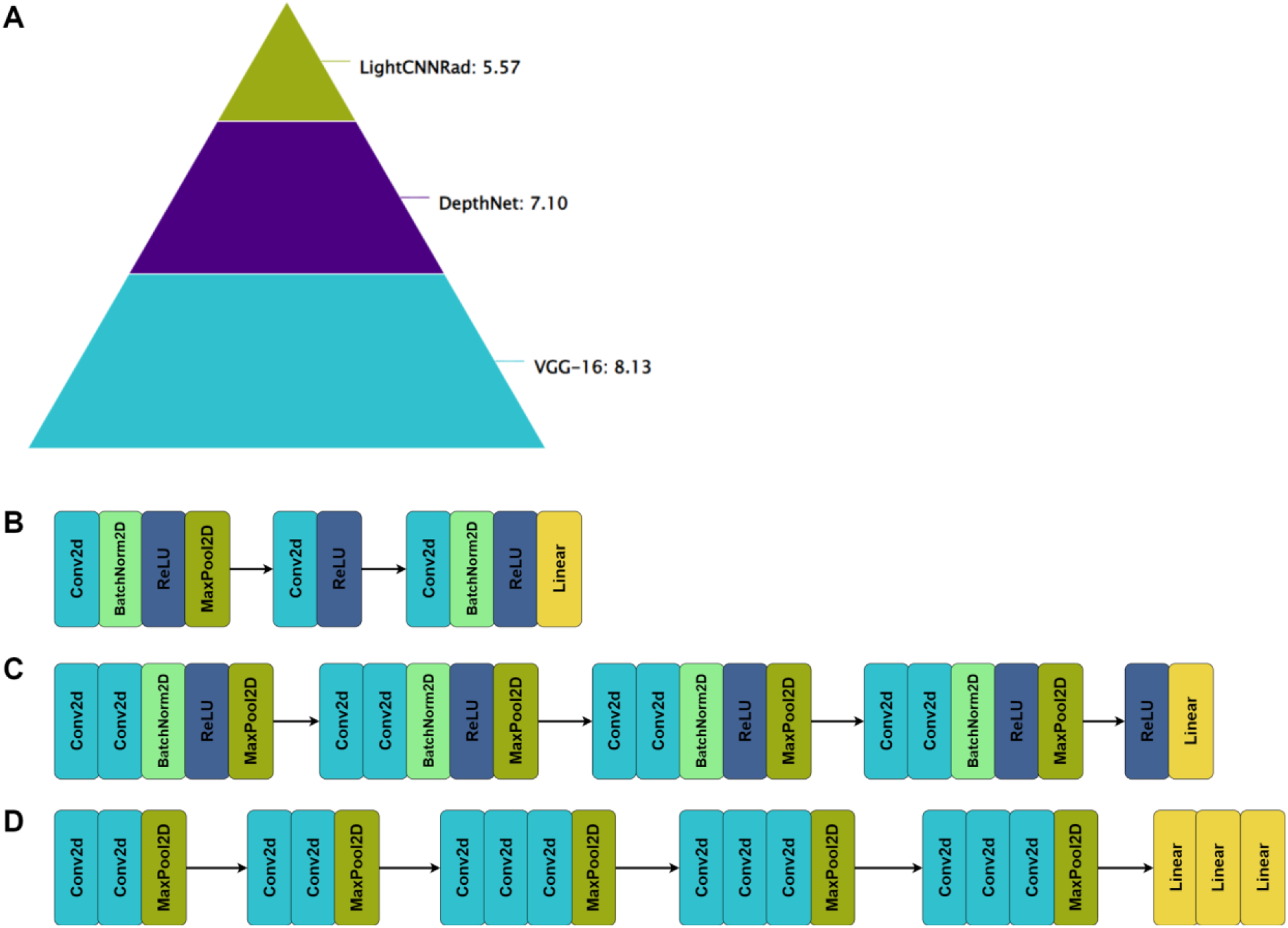
A) Log scale of the number of parameters and the architectures of LightCnnRad, B) DepthNet, and C) VGG-16 deep learning models.

### Transfer Learning

Transfer learning involves utilizing a pre-trained model for new classification and prediction tasks, leveraging its prior training on extensive datasets to enhance performance on related but distinct problems. This approach offers several advantages, such as reducing the training time and providing good levels of generalizability due to the model’s exposure to a vast array of data during its initial training. However, transfer learning also has limitations. Pre-trained models often embed general features from the images they were trained on, which may not be optimal for specific contexts, such as medical imaging, where the features of interest can be quite distinct [26].

VGG-16 is one of the most renowned deep learning architectures used in transfer learning, initially trained on the ImageNet dataset, which contains over 14 million high-resolution images across 1000 different categories [24]. VGG-16 is characterized by its depth, consisting of 16 layers—13 convolutional layers and 3 fully connected layers— demonstrating remarkable performance across various applications [27,28]. To compare the robustness and efficiency of our proposed models, LightCnnRad and DepthNet, with a well-established model, we conducted a comparative analysis involving VGG-16. Using PyTorch, we implemented VGG-16 and set the learning rate to 0.001 to ensure consistency with our models’ training. We applied two approaches: fine-tuning the pre-trained VGG-16 with all the datasets and training it from scratch on the same datasets.

Fine-tuning involves adjusting the weights of the pre-trained VGG-16 model with our specific datasets, allowing the model to adapt its learned features to the new data while retaining the beneficial features learned from the original ImageNet dataset [29]. This method can significantly speed up training and improve performance, especially when the new dataset is limited in size [30]. Training from scratch, however, involves initializing the model’s weights randomly and training it entirely on our datasets without leveraging prior knowledge from ImageNet, providing a baseline to assess the benefits of transfer learning [30]. This comparative analysis aimed to evaluate whether our simplified architectures, LightCnnRad and DepthNet, can achieve similar or superior performance to the traditional and more complex VGG-16 model, but with reduced computational complexity. For VGG- 16, we employed both transfer learning and training from scratch to fully explore its capabilities. In contrast, due to the lightweight nature of LightCnnRad and DepthNet, we focused solely on training from scratch to assess their performance.

## Data Availability

All data links produced in the present study are available in Table 1 of this paper. Due to privacy and security concerns, as well as the sensitivity of medical data, the MR images from the prostate dataset analyzed in this study are not publicly available. The complete codebase and detailed architecture of the algorithms, along with step-by-step instructions, are accessible in our GitHub repository at https://github.com/PKhosravi-CityTech/LightCNNRad-DepthNet.

## Results

### Overall Performance Comparison

Our experimental results indicate that LightCnnRad (mean AUC = 0.82) and DepthNet (mean AUC = 0.85) achieve performance comparable to the pre-trained VGG-16 model (mean AUC = 0.80). Notably, all of these algorithms outperform the VGG-16 model trained from scratch (mean AUC = 0.74). Table 2 summarizes the performance of all four algorithms, detailing the mean AUC, accuracy (ACC), sensitivity, and specificity, along with their respective standard deviations. The confusion matrices corresponding to the four models are provided in Figures 2-5 in the Supplementary Material. Among the models, DepthNet exhibits the highest mean accuracy (ACC) at 0.81, followed closely by LightCnnRad and the pre-trained VGG-16. Additionally, DepthNet shows superior mean specificity (0.87) compared to the pre-trained VGG-16 (0.81), though it has a slightly lower sensitivity (DepthNet: 0.71, VGG-16: 0.74), underscoring its effectiveness in accurately identifying negative cases.

**Table 2.** Overall performance of DepthNet, LightCnnRad, pre-trained VGG-16, and trained from scratch VGG-16 across various metrics.

| Algorithm | Mean AUC | Std Dev AUC | Mean ACC | Std Dev ACC | Mean Sensitivity | Std Dev Sensitivity | Mean Specificity | Std Dev Specificity |
| --- | --- | --- | --- | --- | --- | --- | --- | --- |
| Depthnet | 0.85 | 0.15 | 0.81 | 0.12 | 0.71 | 0.25 | 0.87 | 0.08 |
| LightCnnRad | 0.82 | 0.13 | 0.76 | 0.15 | 0.66 | 0.33 | 0.82 | 0.20 |
| Pretrained VGG-16 | 0.80 | 0.24 | 0.78 | 0.21 | 0.74 | 0.28 | 0.81 | 0.25 |
| Scratch VGG-16 | 0.74 | 0.21 | 0.72 | 0.18 | 0.62 | 0.32 | 0.79 | 0.18 |

We employed the bootstrapping method to estimate confidence intervals for the differences in AUC scores among the algorithms. This non-parametric technique involves repeatedly resampling the dataset to generate simulated samples, thereby providing a more robust measure of variability, particularly beneficial for complex models or when dealing with small sample sizes (refer to Figure 6 in the Supplementary Material). The standard deviations presented in Table 2 for the AUC, ACC, sensitivity, and specificity metrics were calculated using the Numpy package in Python, which underscores the performance variability across the models. Notably, LightCnnRad exhibited lower standard deviations (0.13 for AUC and 0.15 for ACC) alongside DepthNet (0.15 for AUC and 0.13 for ACC), indicating greater outcome stability when compared to the pre-trained VGG-16 (0.24 for AUC and 0.21 for ACC) and the VGG-16 trained from scratch (0.21 for AUC and 0.18 for ACC). This methodological approach facilitates a rigorous evaluation of model performance while effectively accounting for data uncertainties.

The following sections provide a detailed analysis of the model’s performance on each dataset, highlighting the strengths and weaknesses observed in their classification key metrics.

### CT Datasets

All models demonstrated exceptional performance on the COVID-19 CT dataset. Both LightCnnRad and VGG-16, whether pre-trained or trained from scratch, achieved perfect AUC scores of 1.00, while DepthNet also exhibited strong performance with an AUC of 0.93. This consistently high performance across all models underscores the relative ease of distinguishing COVID-19 in CT images, particularly when a substantial amount of data is available (4000 images per class). The large dataset size likely contributed to the models’ ability to generalize well, enabling accurate and reliable classification of COVID- 19 cases (Figure 4 and Figure 1 in the Supplementary Material).

**Figure 4.**
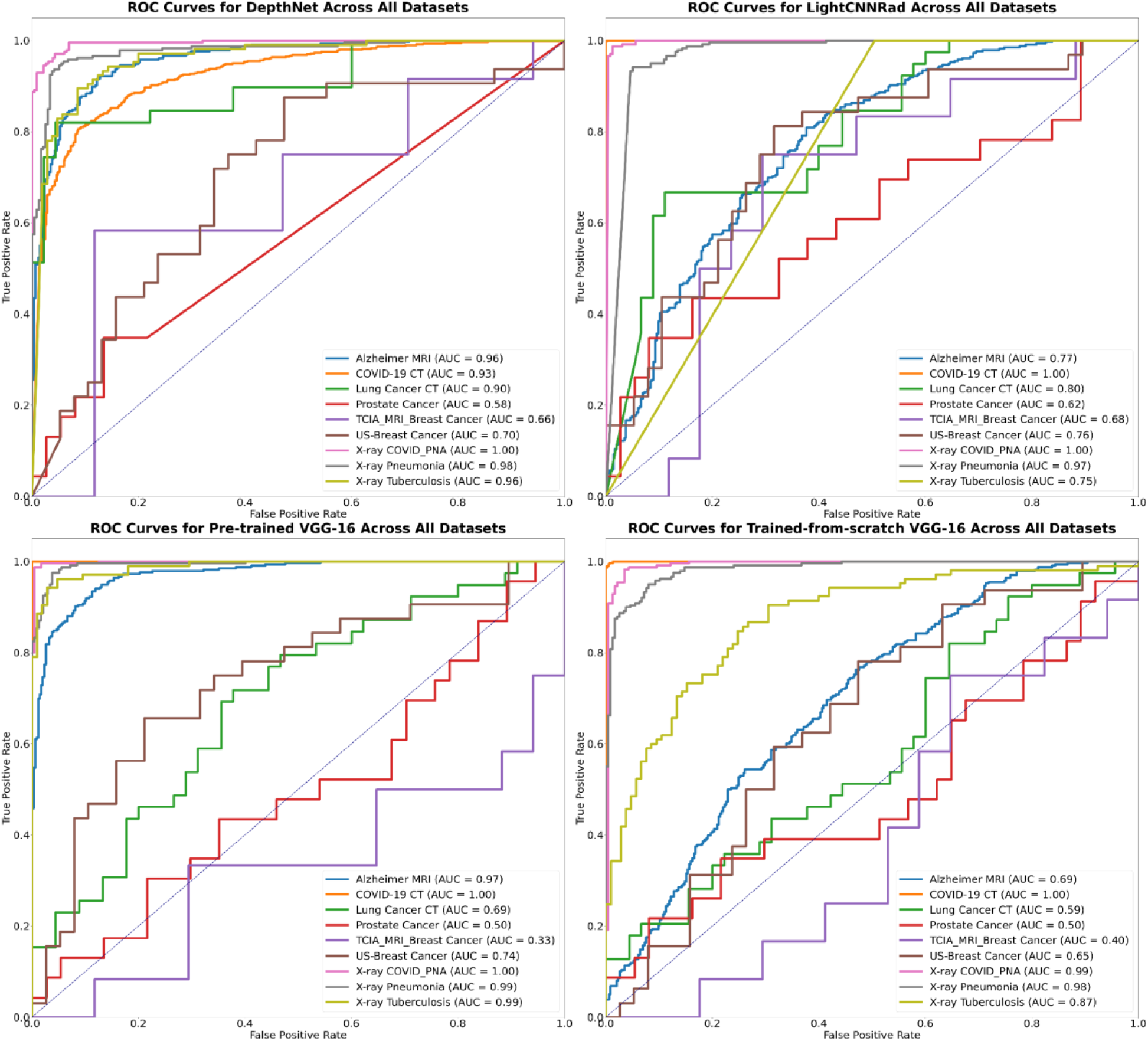
Comparison of AUC Scores for DepthNet, LightCnnRad, and VGG-16 Models.

**Figure 5.**
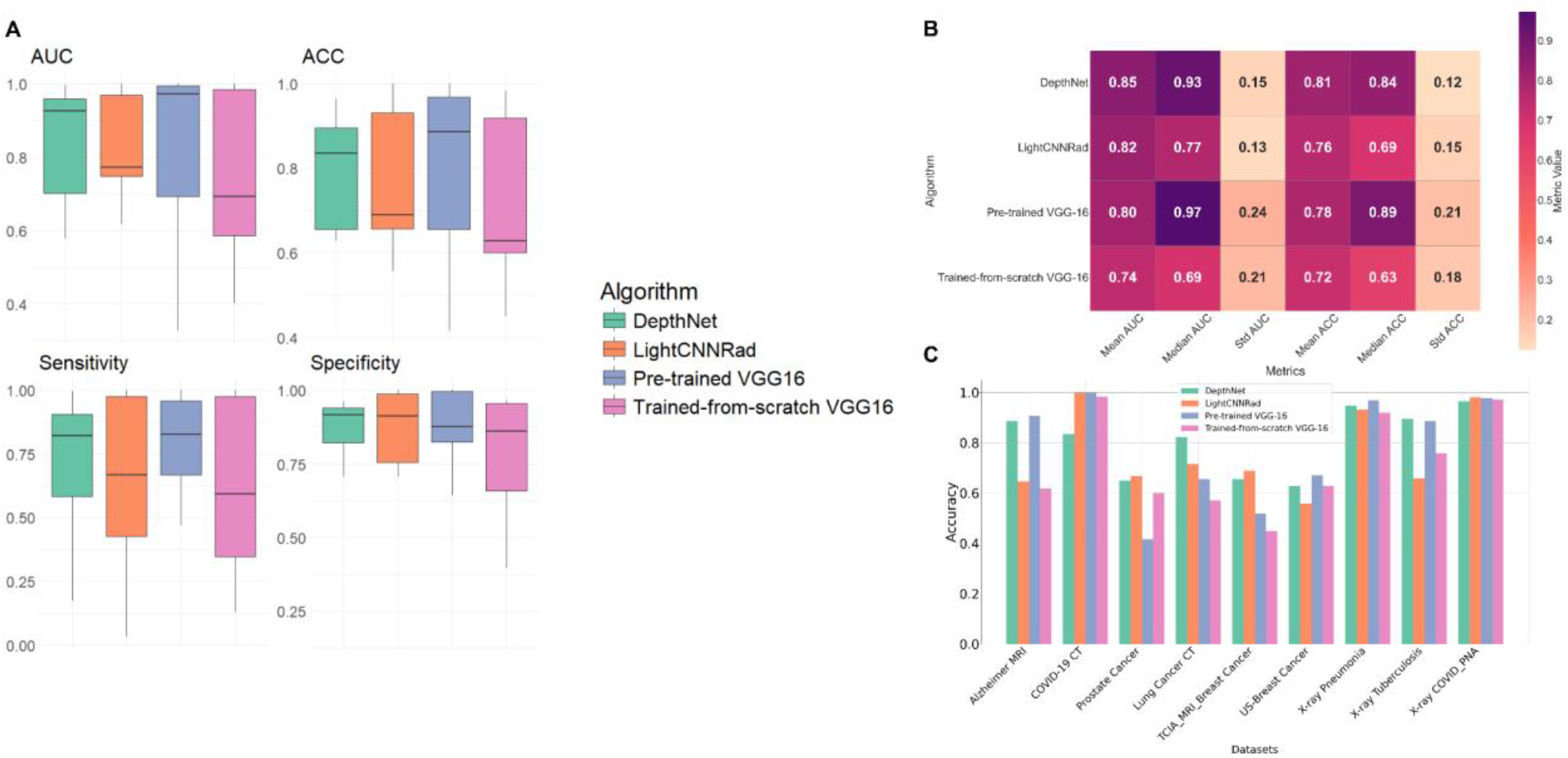
(A) Box plots of performance metrics for different algorithms. (B) Heatmap of mean metrics for different algorithms. (C) Accuracy comparison of DepthNet, LightCnnRad, and VGG-16 models.

DepthNet (AUC = 0.90) and LightCnnRad (AUC = 0.80) outperformed both the pre-trained VGG-16 (AUC = 0.69) and VGG-16 trained from scratch (AUC = 0.59) on the lung cancer CT dataset, which consisted of fewer than 300 images per class. This performance disparity is likely due to the small dataset size (Figure 1A), which tends to favor models with lower complexity. The lightweight architectures of DepthNet and LightCnnRad make them less susceptible to overfitting, allowing them to deliver more stable and reliable results on limited data. These findings emphasize the value of streamlined architectures in data-constrained environments, making them particularly effective for tasks like lung cancer CT image classification (Figure 4 and Figure 1 in the Supplementary Material).

The results highlight the critical importance of selecting model architectures that align with the size and nature of the available data, particularly in clinical contexts where data may be scarce.

### MRI Datasets

For the Alzheimer’s MRI dataset, DepthNet (AUC = 0.96) and pre-trained VGG-16 (AUC = 0.97) exhibit excellent performance, indicating their robustness in detecting Alzheimer’s disease through MRI scans. LightCnnRad (AUC = 0.77) and VGG-16 trained from scratch (AUC = 0.69) show comparatively lower performance. The medium-sized dataset, with around 2000 images per class (Table 1), is sufficient for more complex models like pre-trained VGG-16 and DepthNet to perform well. However, VGG-16 trained from scratch struggles due to insufficient data to learn effectively. LightCnnRad’s simpler architecture, while beneficial for smaller datasets, appears to be less effective for this medium-sized dataset, suggesting it might require further optimization or additional training data to improve its performance (Figure 4 and Figure 1 in the Supplementary Material).

LightCnnRad (AUC = 0.68) outperformed DepthNet (AUC = 0.66), pre-trained VGG-16 (AUC = 0.33), and VGG-16 trained from scratch (AUC = 0.40) in predicting neoadjuvant therapy outcomes using the multi-sequence TCIA-Breast Cancer MRI dataset. This suggests that LightCnnRad is more effective at capturing the subtle features necessary for this challenging classification task. Despite incorporating both pre- and post-contrast MRI images to leverage multi-sequence data, the overall model performance remained modest. The complexity added by using multi-sequence images, combined with the relatively small number of patient cases (188 patients) despite the high number of images per class (about 10,000 images per class), likely contributed to the difficulty faced by the more complex models like VGG-16 in achieving high accuracy. The complexity of this task is further heightened by the challenge of predicting pathology based on treatment outcomes without targeted tumor cropping, a limitation stemming from the absence of expert radiologists during preprocessing. LightCnnRad’s simpler architecture, however, allowed it to navigate these complexities more effectively, avoiding overfitting and better managing the intricacies of the data. These findings underscore the importance of model selection in scenarios involving high task complexity and limited patient numbers, where streamlined models like LightCnnRad can offer a performance advantage over more complex architectures (Figure 4 and Figure 1 in the Supplementary Material).

LightCnnRad outperformed all other models on the Prostate Cancer MRI dataset, achieving an AUC of 0.62. DepthNet (AUC = 0.58) also performed better than both the pre-trained VGG-16 (AUC = 0.50) and the VGG-16 model trained from scratch (AUC = 0.50). The relatively low AUC scores for the VGG-16 models suggest that these more complex architectures struggled to classify the prostate cancer MRI images effectively, performing no better than random chance. This result highlights the difficulty these models faced in capturing the subtle and intricate patterns present in the prostate MRI images. Even though the pre-trained VGG-16 model was trained on large and diverse datasets such as ImageNet, these datasets are not optimized for medical imaging tasks like prostate cancer detection, which may explain the lackluster performance. In contrast, LightCnnRad demonstrated superior performance, likely due to its streamlined architecture, which appears better suited to the specific features present in the preprocessed prostate MRI images. The preprocessing steps—such as cropping the images to focus on the prostate gland and the most relevant anatomical structures—likely contributed to the improved performance of the simpler models. It’s also worth noting that prostate cancer aggressivity correlates well with the PIRADS radiological classification system. PIRADS considers multiple imaging modalities, including diffusion-weighted imaging (DWI), which was not used or trained in this study. Our results suggest that, although classifying prostate cancer from MRI images remains challenging, using simpler and specialized models like DepthNet and LightCnnRad, combined with effective preprocessing techniques, can improve performance. Future work could further explore the inclusion of additional imaging modalities, such as DWI, to enhance model accuracy and better reflect the comprehensive PIRADS classification.

This highlights the importance of selecting the appropriate model architecture and preprocessing techniques, particularly in complex medical imaging tasks where data characteristics differ significantly from more general datasets (Figure 4 and Figure 1 in the Supplementary Material).

### Ultrasound Dataset

For the US-Breast Cancer dataset (200 images per class), the performance of the models aligns with expectations: lighter models tend to perform better with limited data. LightCnnRad achieved the highest AUC at 0.76, underscoring its effectiveness in interpreting ultrasound images in such scenarios. DepthNet (AUC = 0.70) and pre-trained VGG-16 (AUC = 0.74) also performed well, indicating their capability to handle ultrasound images, though they might benefit from further optimization. In contrast, VGG-16 trained from scratch showed the lowest performance with an AUC of 0.65, likely due to the model’s complexity and the insufficient available data for effective training from scratch. These results suggest that while complex models can benefit from transfer learning, simpler architectures like LightCnnRad can offer superior performance in contexts with limited data and high task complexity. This finding emphasizes the importance of selecting appropriate model architectures based on the data available and the specific challenges of the task (Figure 4 and Figure 1 in the Supplementary Material).

### X-ray Datasets

The X-ray datasets used in this study contained a moderate to low number of images (Figure 1), yet all models performed well on these tasks. On the X-ray COVID-19 PNA dataset, all models—LightCnnRad, pre-trained VGG-16, DepthNet, and VGG-16 trained from scratch—achieved perfect AUC scores of 1.00. This exceptional performance indicates that COVID-19 features in X-ray images are relatively straightforward for these models to detect. Similarly, the models performed strongly on the X-ray Pneumonia dataset, with DepthNet (AUC = 0.98), LightCnnRad (AUC = 0.97), pre-trained VGG-16 (AUC = 0.99), and VGG-16 trained from scratch (AUC = 0.98) all showing consistently high AUC values. These results suggest that pneumonia features in X-ray images are effectively captured by all models.

For the X-ray Tuberculosis dataset, the models continued to perform well, though with more variation in their results. Pre-trained VGG-16 achieved the highest AUC at 0.99, followed by DepthNet (AUC = 0.96), VGG-16 trained from scratch (AUC = 0.87), and LightCnnRad (AUC = 0.75). The superior performance of the pre-trained VGG-16 suggests that pre-training on large, diverse datasets may provide an advantage in this application, while LightCnnRad’s lower AUC indicates that it may be less effective in this particular task (Figure 4 and Figure 1 in the Supplementary Material).

### Interpretation beyond algorithms

The findings from this study highlight the critical importance of selecting appropriate model architectures based on the specific characteristics of the dataset and the clinical task at hand. The consistent performance of DepthNet and LightCNNRad, particularly in scenarios with limited data, underscores the advantage of streamlined architectures in medical image analysis. These models demonstrate that, in many cases, simpler architectures can achieve or even exceed the performance of more complex models like VGG-16, especially when the available data is not extensive (Figure 5).

The results suggest that while VGG-16—especially in its pre-trained form—generally offers robust performance across various tasks, its complexity can be a disadvantage when the dataset size is constrained or when the task requires the model to discern subtle patterns from a limited number of examples. In contrast, the simpler architectures of DepthNet and LightCnnRad are less prone to overfitting and can better generalize under these conditions. This finding suggests that increasing model complexity does not necessarily result in better performance in medical image analysis.

Moreover, the study underscores the critical role of preprocessing techniques in enhancing model performance. For instance, the application of cropping and other preprocessing steps helped to focus the analysis on the most relevant areas of the images, which was particularly beneficial for simpler models like LightCNNRad and DepthNet. This indicates that thorough preprocessing can reduce the need for more complex models by delivering cleaner and more focused data for analysis.

The variability in performance observed across different models and datasets also highlights the importance of understanding the underlying data characteristics. For example, the high variability in VGG-16’s performance across different tasks suggests that while it is a powerful model, its effectiveness can be highly dependent on the nature of the data and the specific task. This reinforces the need for a multifactorial approach in model selection, where factors such as dataset size, image quality, and the specific clinical question are all considered.

Overall, these findings advocate for a more nuanced approach to model selection in medical imaging, one that considers not just the algorithmic complexity but also the practical aspects of the data and the task. By focusing on the broader context in which these models are applied, we can develop more effective, reliable, and clinically relevant tools for medical image analysis. This holistic perspective is essential for advancing the field and ensuring that the models developed are not only accurate but also generalizable across different clinical settings.

## Discussion

The development of LightCnnRad and DepthNet represents a significant advancement in the application of CNNs to radiological image analysis. Our study aimed to address the limitations of traditional deep learning models, such as VGG-16, by proposing simpler architectures that maintain high performance while reducing computational complexity.

The experimental results demonstrate that LightCnnRad (AUC = 0.82) and DepthNet (AUC = 0.85) achieve accuracy rates comparable to the pre-trained VGG-16 model (AUC = 0.80) across a validation set of diverse radiological images. Despite their simpler architectures, both models exhibited strong classification performance across various imaging modalities, including MRI, CT, X-ray, and Ultrasound. This highlights their robustness and generalizability in medical image analysis.

One of the key advantages of LightCnnRad and DepthNet is their computational efficiency. The reduced number of parameters in these models translates to faster training times and lower computational resource requirements (Figures 6 to 9 in the Supplementary Material). This is particularly beneficial in clinical settings where real-time processing and analysis are crucial. For example, LightCnnRad, with its simpler architecture, maintains high accuracy while significantly cutting down on the training duration compared to more complex models. DepthNet, leveraging depthwise separable convolutions, achieves a similar reduction in computational demands without compromising performance.

Our comparative analysis with the VGG-16 model, both pre-trained and trained from scratch, underscores the potential of simplified CNN architectures. While the pre-trained VGG-16 benefits from transfer learning due to its extensive pre-training on the ImageNet dataset, our models, designed specifically for radiological applications, can adapt efficiently to the nuances of medical imaging data. This adaptability is crucial for tasks that require specialized feature extraction, such as distinguishing between benign and malignant tumors or classifying different stages of diseases.

The results from fine-tuning VGG-16 also highlight a critical insight: pre-trained models, while powerful, may not always provide optimal performance for domain-specific tasks. The features learned from general datasets like ImageNet may not align perfectly with the features needed for medical image analysis. Our study shows that tailored architectures, designed with domain-specific requirements in mind, can achieve competitive or even superior performance with less computational overhead, which is in alignment with previous findings that emphasize the benefits of using pre-trained models on medical image datasets over those trained on natural images like ImageNet [31].

The implications of our findings for clinical practice are significant. By utilizing models like LightCnnRad and DepthNet, healthcare providers can implement efficient, real-time analysis of radiological images without the need for extensive computational infrastructure. This can facilitate faster diagnosis and treatment decisions, ultimately improving patient outcomes.

Future work will focus on further optimizing these models and validating their performance in real-world clinical settings. This includes expanding the datasets to encompass a wider range of imaging modalities and conditions, as well as exploring the integration of these models into existing diagnostic workflows. Additionally, we plan to investigate the potential of combining our simplified architectures with other machine learning techniques, such as ensemble learning, to further enhance performance.

In summary, our study challenges the common misconception that more complex architectures always lead to higher performance. The high performance of LightCnnRad and DepthNet across various tasks suggests that simpler models can be highly effective, especially when carefully designed for specific applications. The findings emphasize the importance of a multifactorial approach in developing and applying CNNs to radiological image analysis, ensuring that models are not only accurate but also computationally efficient and adaptable to diverse clinical environments.

## Conclusion

This study introduced LightCnnRad and DepthNet, two novel CNN architectures specifically designed for radiological image analysis. These models achieved performance comparable to the more complex pre-trained VGG-16, while significantly reducing computational complexity and training times. LightCnnRad and DepthNet excelled across various imaging modalities, including MRI, CT, X-ray, and Ultrasound, proving their versatility and efficiency in medical imaging tasks.

Our findings underscore that simpler architectures can achieve competitive performance, highlighting the potential of models like LightCnnRad and DepthNet in clinical settings where computational resources and real-time processing are critical. Moreover, our analysis goes beyond algorithmic performance, emphasizing the importance of a holistic approach that includes preprocessing techniques and consideration of external factors to enhance model effectiveness.

Future research will focus on optimizing these models further and validating their performance in real-world clinical environments. In conclusion, LightCnnRad and DepthNet represent promising tools for efficient and accurate radiological image analysis, advancing the capabilities of CNNs in medical imaging by combining high performance with computational efficiency.

## Data Availability

All data produced in the present study are available upon reasonable request to the authors

https://github.com/PKhosravi-CityTech/LightCNNRad-DepthNet

## Acknowledgments

We would like to thank the BioMind AI Lab (https://sites.google.com/view/biomind-ai-lab) for their invaluable support and contributions to this research. This work would not have been possible without the dedication and hard work of our students and research assistants.

## Contributions

S.M. led the study, contributed to developing the models, performed the data analysis, and wrote the manuscript. P.K. developed the models, contributed to writing the manuscript, and supervised the entire study. A.S.M. assisted S.M. in preparing the primary dataset organization, downloading public datasets, and contributed to the early stages of data analysis. S.S. prepared the in-house data and assisted in the interpretation of the results. D.R., W.L.H., R.G., T.L., E.H., and A.H. contributed to the application of the models on various datasets. J.Z., I.D., and V.P. provided supervision, guidance to the students, and critical revisions to the manuscript. All authors participated in reviewing, editing, and approving the final version of the manuscript.

## Supplementary Material

**Figure S1.**
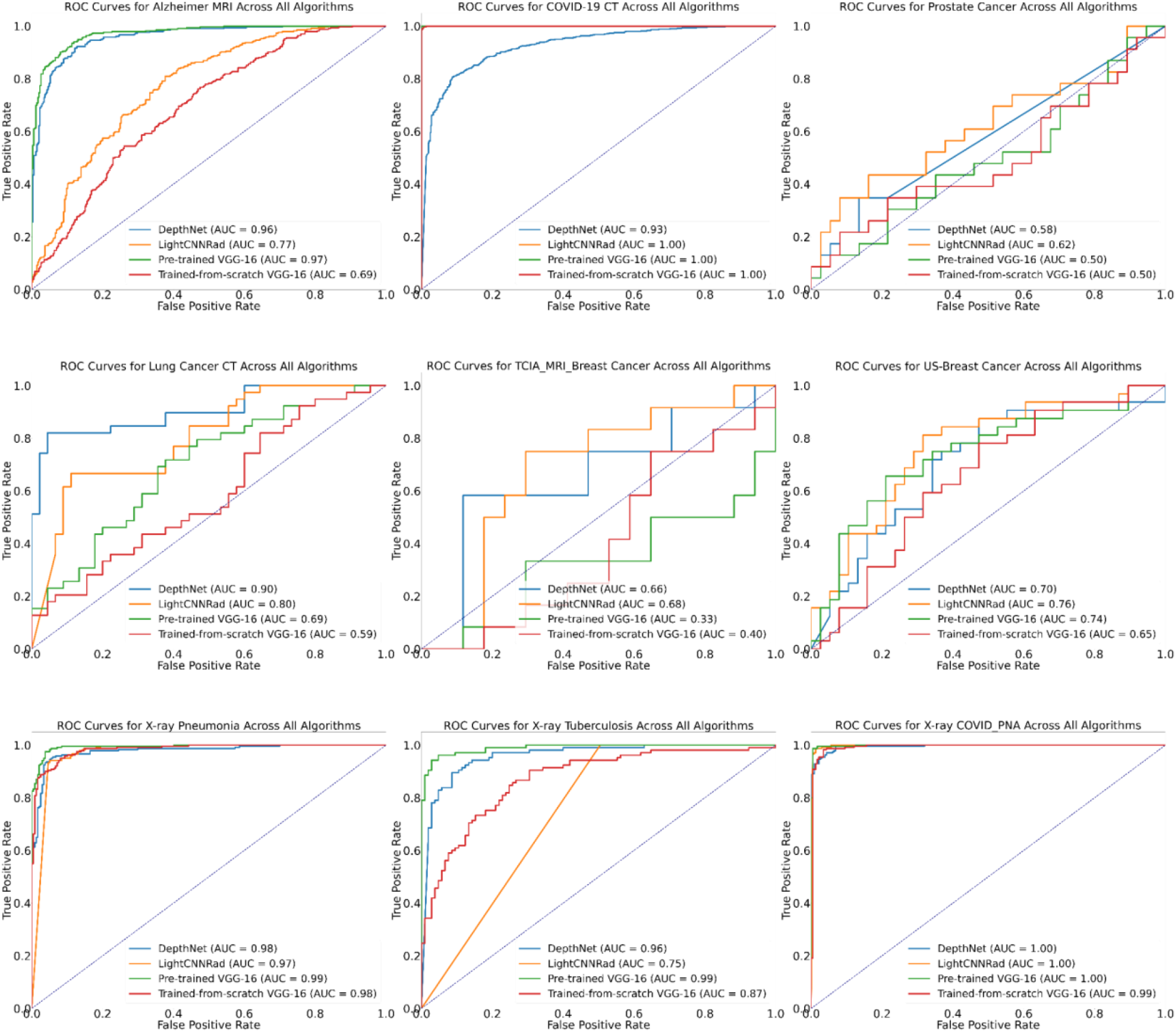
Comparison of AUC values for DepthNet, LightCNNRad, pre-trained VGG-16, and VGG-16 trained from scratch across nine datasets.

**Figure S2.**
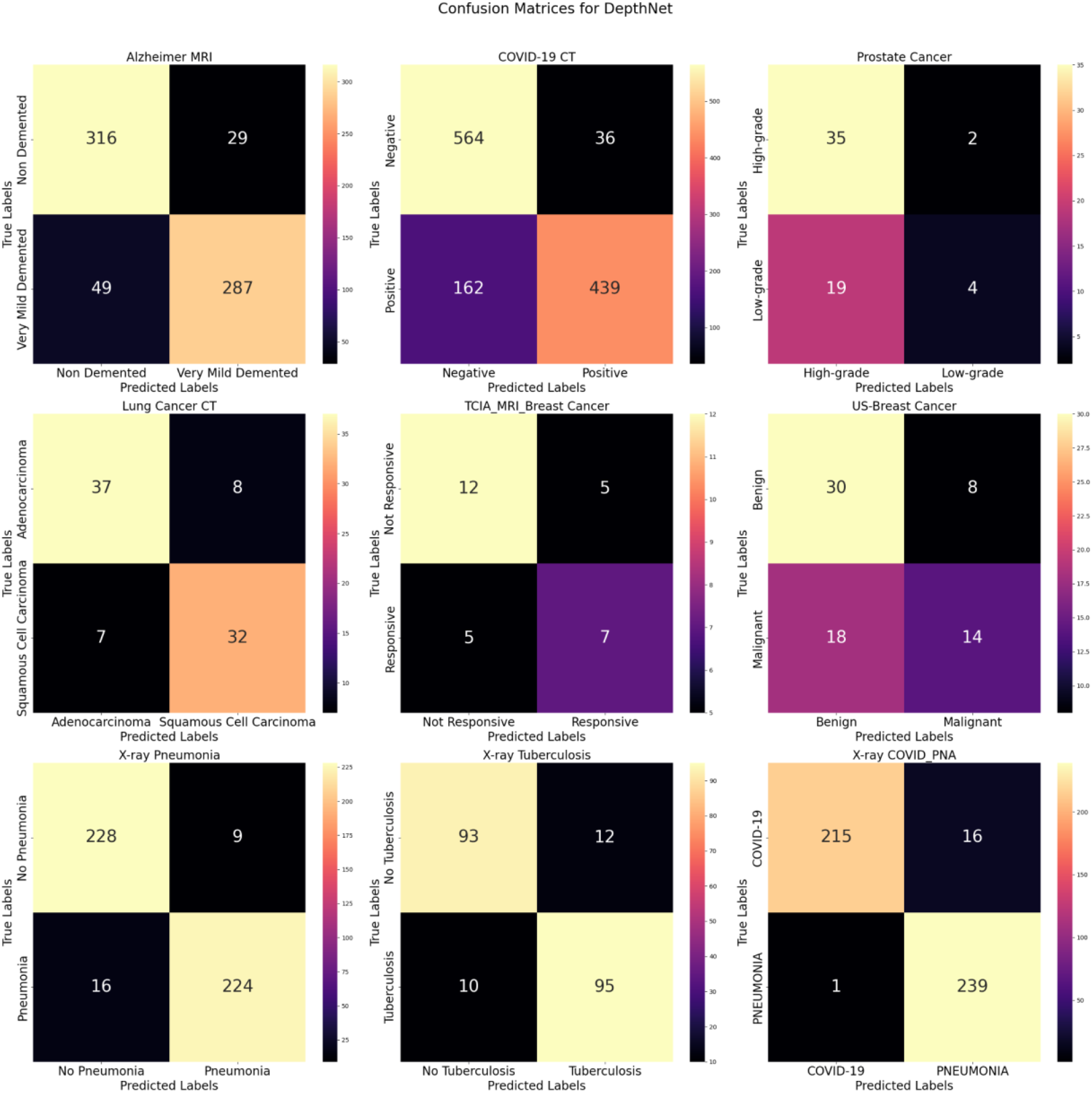
Confusion matrices for DepthNet across all datasets.

**Figure S3.**
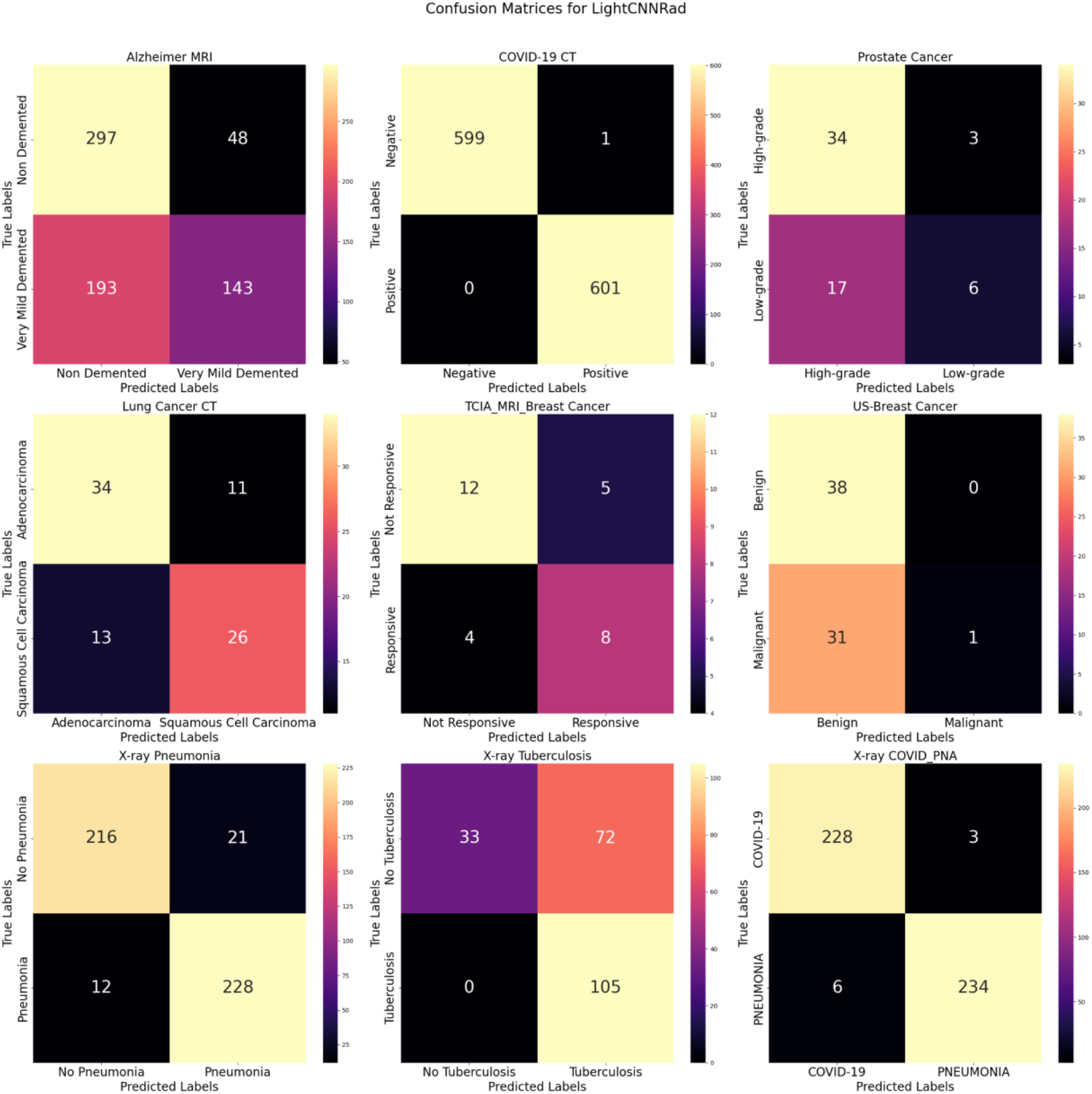
Confusion matrices for LightCNNRad across all datasets.

**Figure S4.**
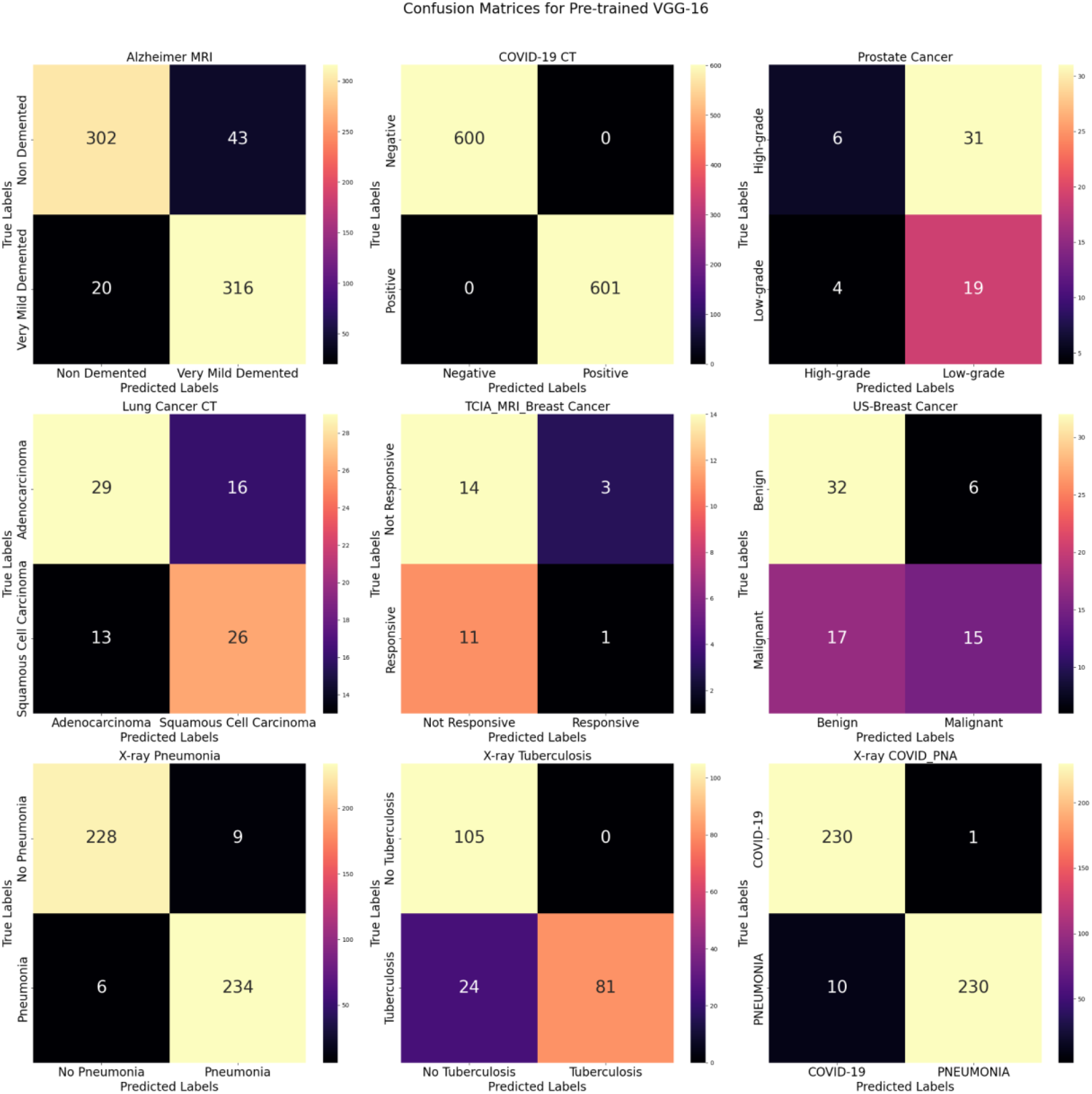
Confusion matrices for the pre-trained VGG-16 across all datasets.

**Figure S5.**
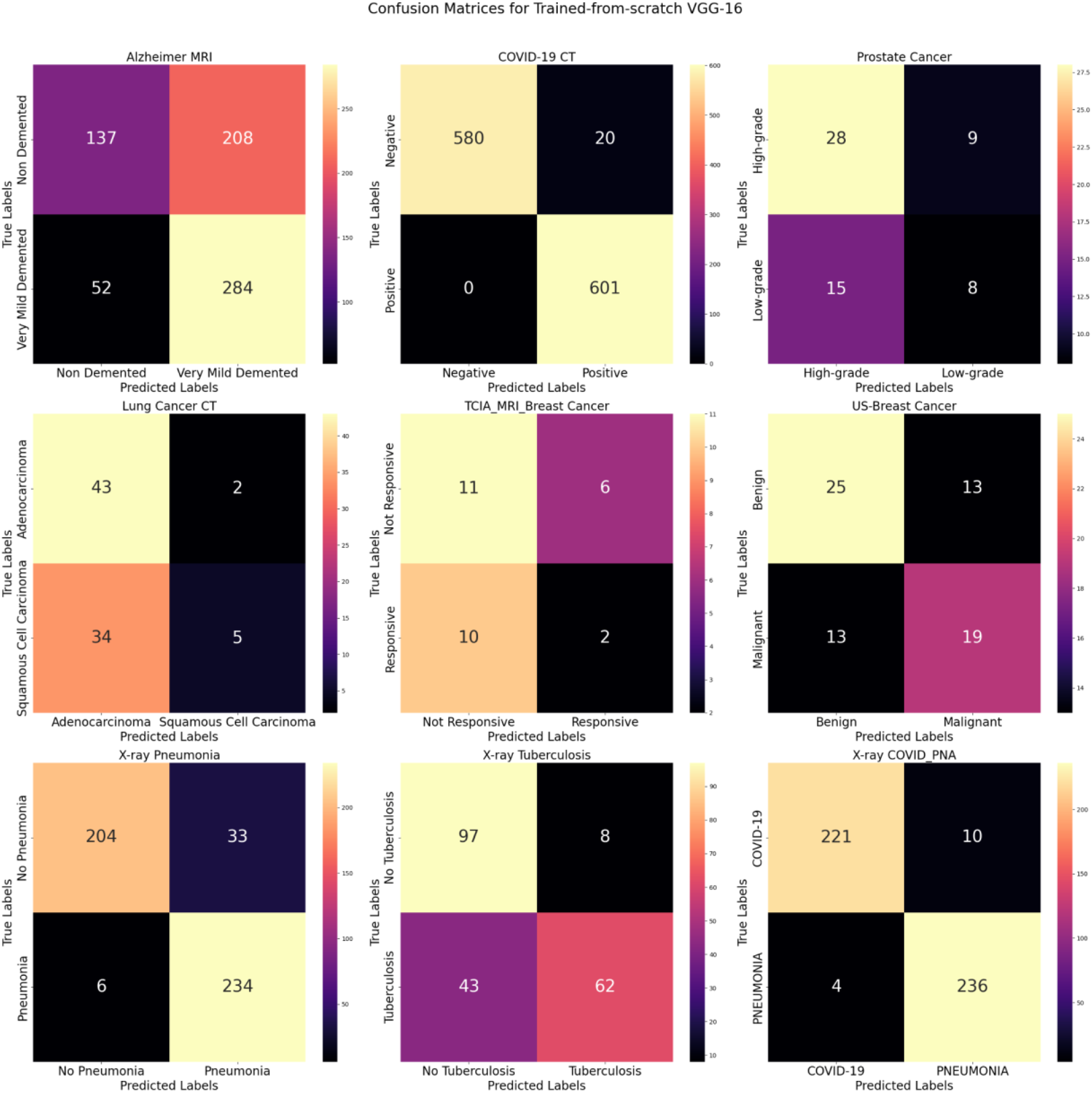
Confusion matrices for the VGG-16 trained from scratch across all datasets.

**Figure S6.**
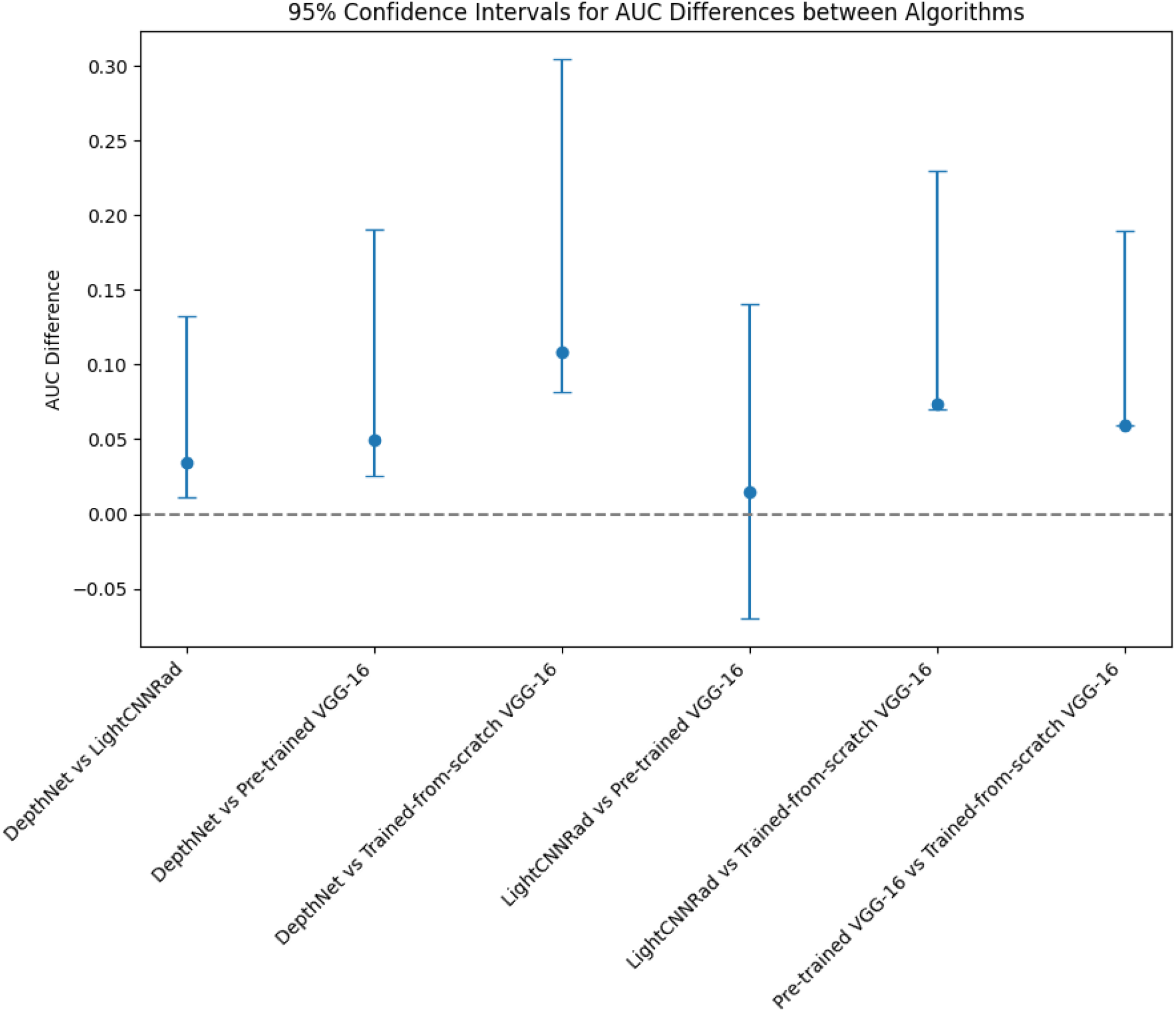
Confidence intervals for the differences in AUC scores between algorithms were estimated using bootstrapping.

**Figure S7.**
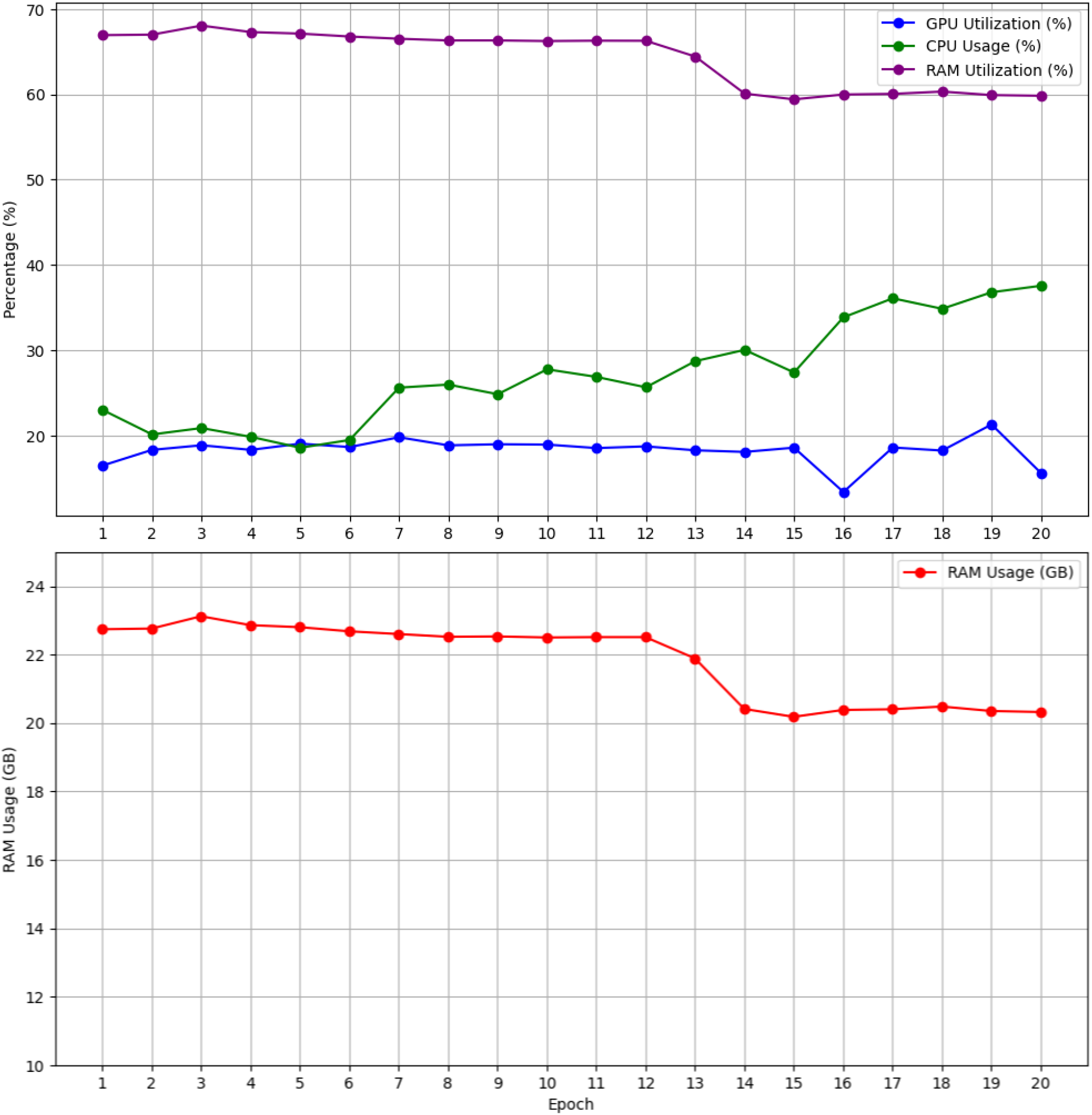
Efficiency of LightCnnRad on the Alzheimer’s MRI dataset, measured by GPU utilization, CPU usage, and RAM utilization.

**Figure S8.**
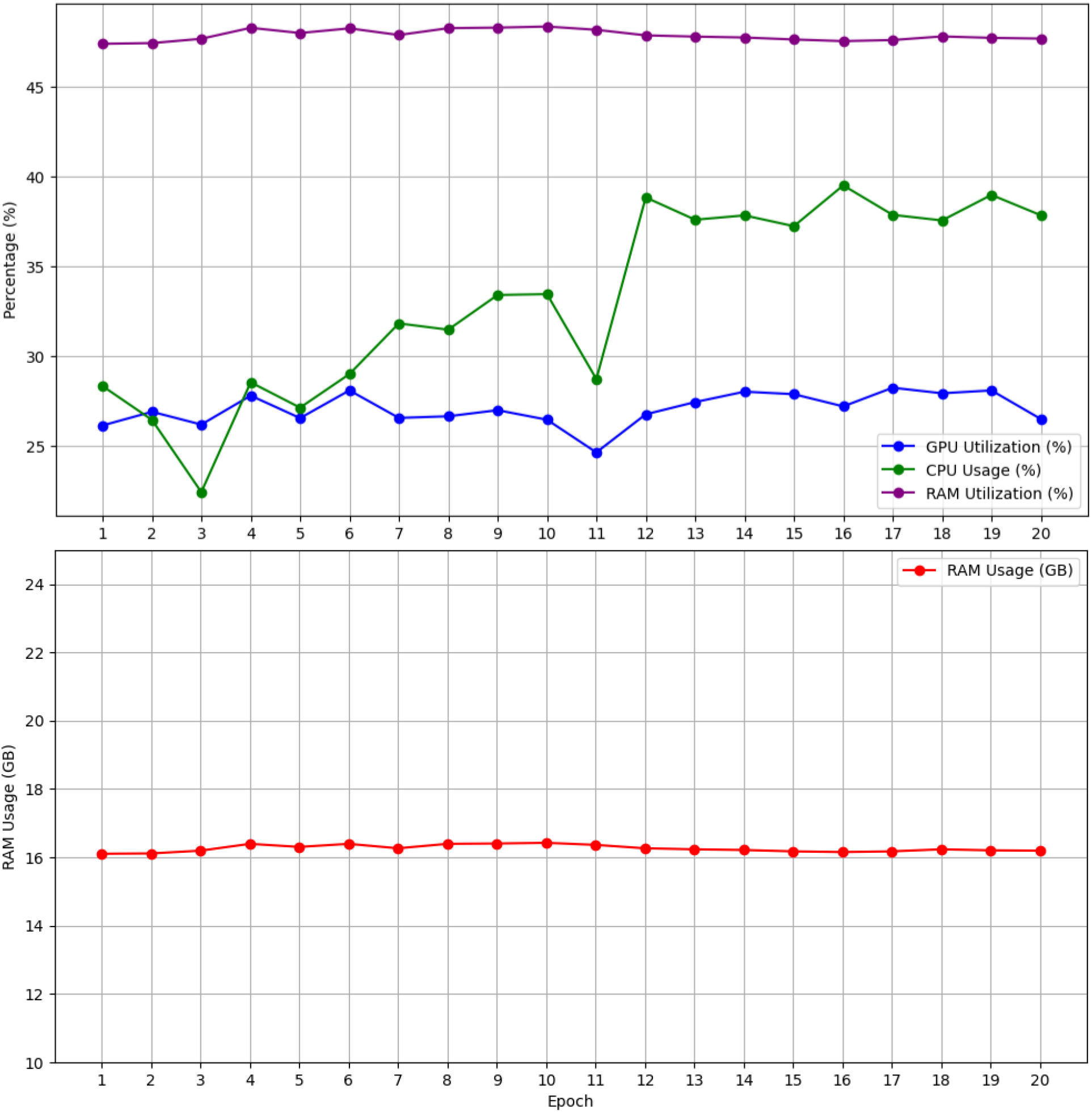
Efficiency of DepthNet on the Alzheimer’s MRI dataset, measured by GPU utilization, CPU usage, and RAM utilization.

**Figure S9.**
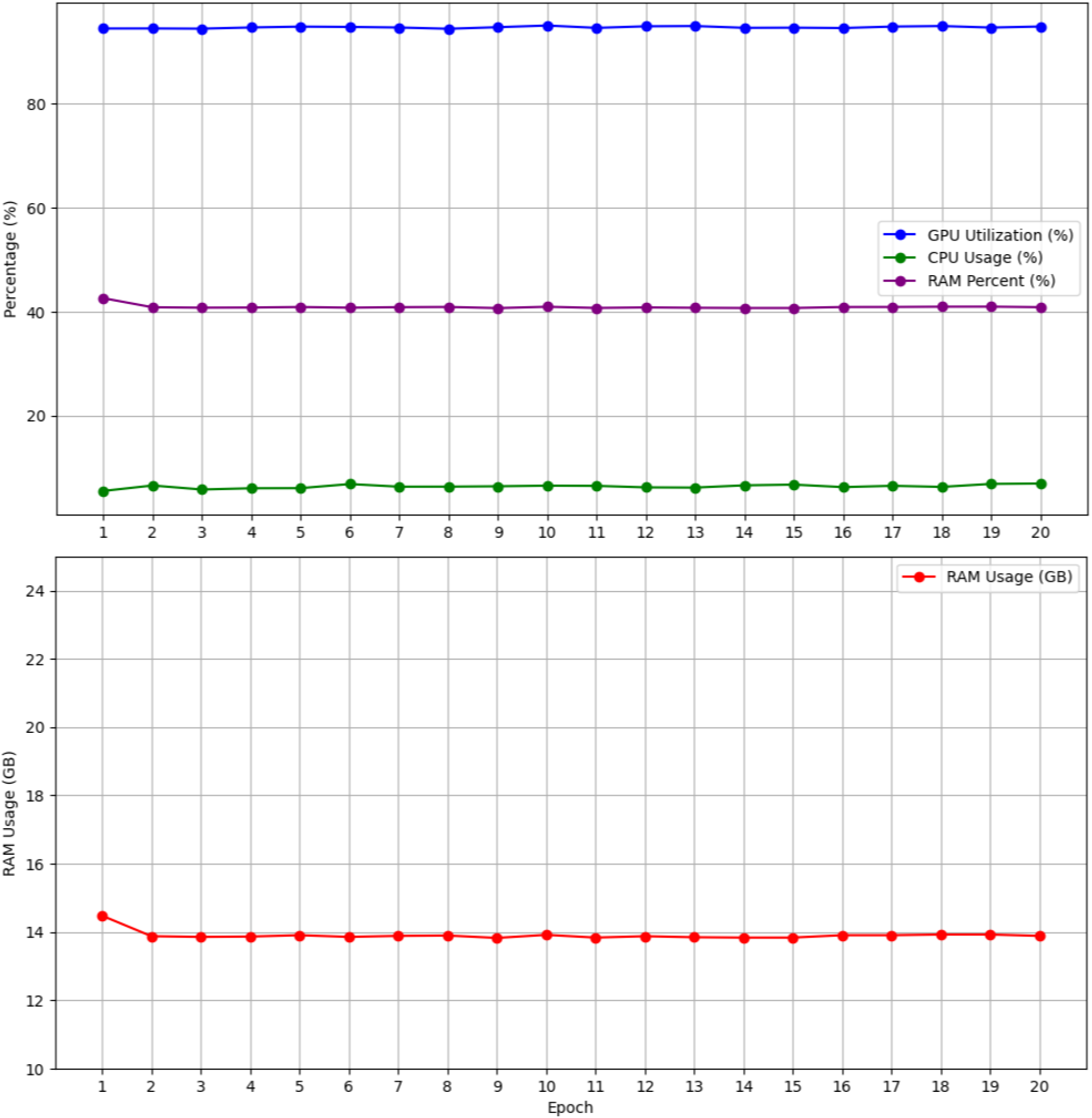
Efficiency of pre-trained VGG-16 on the Alzheimer’s MRI dataset, measured by GPU utilization, CPU usage, and RAM utilization.

**Figure S10.**
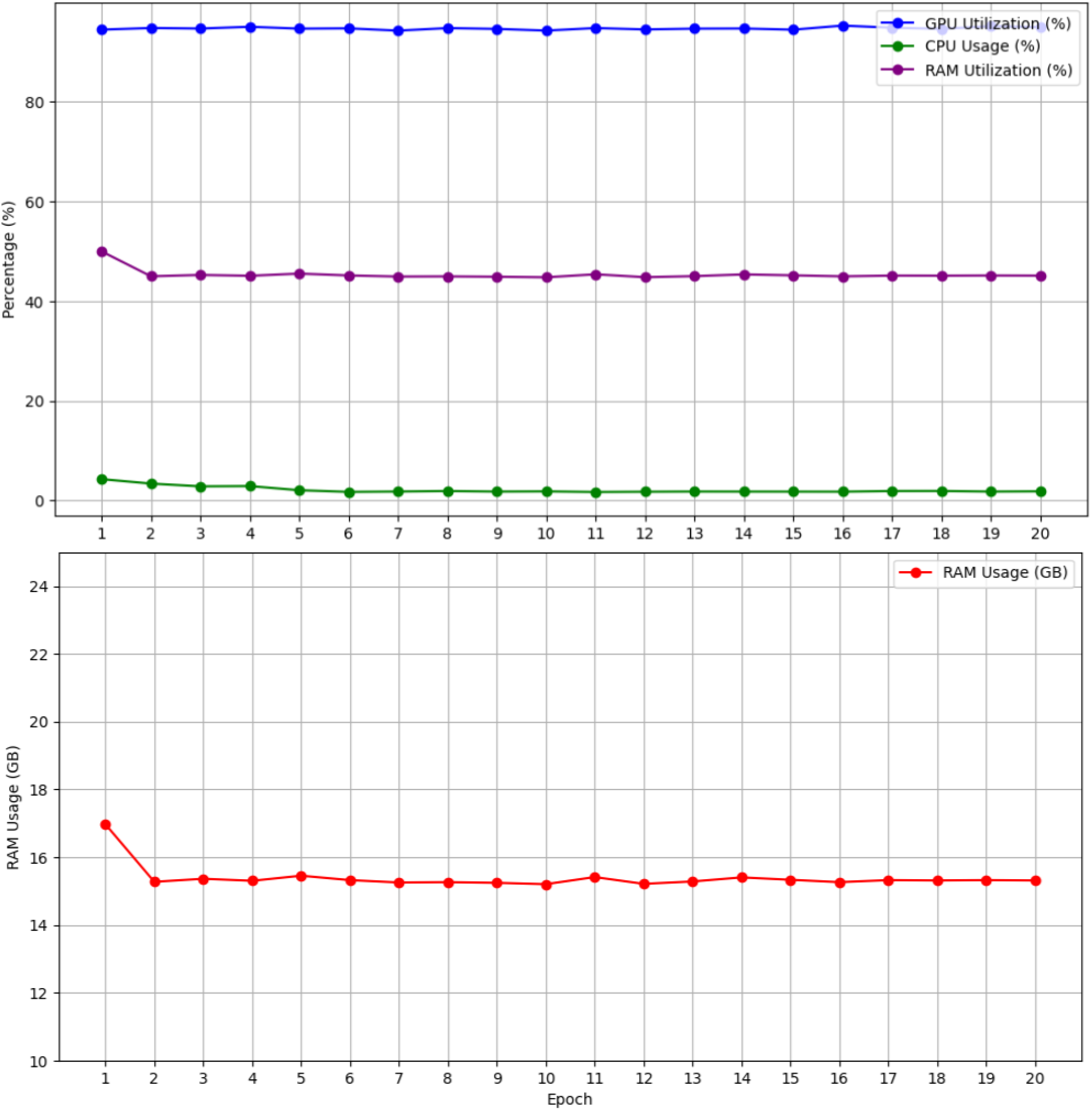
Efficiency of VGG-16 trained from scratch on the Alzheimer’s MRI dataset, measured by GPU utilization, CPU usage, and RAM utilization.

## Notes

### Competing Interest Statement

The authors have declared no competing interest.

### Clinical Protocols

https://github.com/PKhosravi-CityTech/LightCNNRad-DepthNet

### Funding Statement

This study did not receive any funding

### Author Declarations

Ethics committee/IRB of AdventHealth in Orlando, FL, gave ethical approval for this work under protocol number 3009855250.

